# Describing the inputs, activities and outputs of “10,000 Lives”, a coordinated regional smoking cessation initiative in Central Queensland, Australia

**DOI:** 10.1101/2020.09.08.20190264

**Authors:** Arifuzzaman Khan, Kalie Green, Gulam Khandaker, Sheleigh Lawler, Coral Gartner

## Abstract

**Objective:** This study utilised a program logic model to describe the inputs, activities and outputs of the “10,000 Lives” smoking cessation initiative in Central Queensland, Australia

**Design:** A program logic model provided the framework for the process evaluation of “10,000 Lives”. The data were collected through document review, observation and key informant interviews, and subsequently analysed after coding and re-coding into classified themes, inputs, activities and outputs.

**Setting:** The prevalence of smoking is higher in the Central Queensland region of Australia compared to the national and state averages. In 2017, Central Queensland Hospital and Health Services set a target to reduce the percentage of adults who smoke from 16.7% to 9.5% in the Central Queensland region by 2030 as part of their strategic vision (‘Destination 2030’). Achieving this target is equivalent to 20,000 fewer smokers in Central Queensland, which should result in 10,000 fewer premature deaths due to smoking-related diseases. To translate this strategic goal into an actionable smoking-cessation initiative, the “10,000 Lives” health promotion program was officially launched on 1 November 2017.

**Result:** The activities of the initiative coordinated by a senior project officer included building clinical and community taskforces, organising summits and workshops, and regular communications to stakeholders. Public communication strategies (e.g., Facebook, radio, community exhibitions of “10,000 Lives”, and health-related events) were utilised to promote available smoking cessation support to the Central Queensland community.

**Conclusion:** The “10,000 Lives” initiative provides an example of a coordinated health promotion program to increase smoking cessation in a regional area through harnessing existing resources and strategic partnerships (e.g., Quitline). Documenting and describing the process evaluation of the “10,000 Lives” model is important so that it can be replicated in other regional areas with a high prevalence of smoking.

**Strengths and limitations of this study:**

- The study considered a standard evaluation framework (logic model) to describe the program.
- Multiple sources of data were collected and included to describe the process of the program
- The plan for impact evaluation of the program is discussed in the article.
- Some outputs may have been omitted due to lack of systematic documentation of all activities within the project field notes.

## Introduction

Tobacco smoking remains the leading avoidable risk factor that contributes to the burden of death and disease in Australia. The 2015 Australian Burden of Disease Study estimated that 9.3% of the total disease burden, 13.3% of all deaths, and 443,235 Disablity Adjusted Life Years (DALYs) were related to tobacco use.^1,2^ In 2016, twelve percent of the adult population were daily smokers in Australia,^3^ whereas 14.5% of adults in Queensland,^4^ and 16.7% of adults in Central Queensland (CQ) smoked daily.^5^ Reasons for the higher smoking prevalence in CQ may include the higher proportion of the population who experience socioeconomic disadvantage, compared to the state average.^6,7^ Priority populations for smoking cessation assistance identified within CQ include pregnant women (17.0% smoking prevalence),^8^ and people living in some local government areas within CQ, such as Gladstone 19.1% and Rockhampton 17.7% smoking prevalence.^6,7^

Tobacco control and smoking cessation programs are a shared responsibility between the Federal Government and the States and Territories in Australia.^9^ The State and Territory governments implement many tobacco control and smoking cessation programs. Queensland has performed as one of the best states in Australia for tobacco control activities in recent years.^10^ Programs and policies delivered by the Queensland Government include the Quitline service^11^, anti-smoking mass media campaigns, and smoke-free policies and laws. These have contributed to maintaining a downward trend in the daily smoking rate in Queensland over the last few decades.^12^ However, a significantly higher rate of adult smoking than the state average has persisted in some regional areas like the CQ region.^8^

The higher prevalence of smoking in CQ compared with the whole of Queensland led CQ Health and Hospital Service (CQHHS) to prioritise smoking cessation while formulating the region’s strategic health vision (known as ‘Destination 2030’) through a six month consultation process with CQ health personnel, consumers, priority groups, and community partners.^13^ As part of ‘Destination 2030’,^13^ CQHHS set a goal to reduce the adult daily smoking prevalence from 16.7% to 9.5% in CQ by 2030. Accomplishing this goal would be equivalent to 20,000 fewer smokers in CQ which was estimated to result in 10,000 lives that would be saved from premature death due to smoking-related diseases because half of all long-term-smokers die from a smoking related disease.^14^ The strategic goal was translated into an actionable health promotion initiative to increase smoking cessation, which was named “10,000 Lives”.^15^ The name of the “10,000 Lives” initiative builds on the previously highly successful “10,000 Steps Rockhampton” program, which promoted physical activity in Rockhampton.^16,17^

Tobacco control and smoking cessation are priorities of federal and state governments (e.g., Queensland) in Australia, yet, there are always budgets constraints for preventive health promotion.^18^ As such it is imperative to consider how to leverage off existing funded programs with small iterative changes and budgets. A low-cost and locally initiated program like “10,000 Lives” is one such example, where this principle is being applied, with the aim of improving the health and wellbeing of the community. This paper documents the process evaluation of “10,000 Lives” so that researchers, health professionals and policy makers can use this information for future program planning.

## Aim

This study aims to describe the inputs (planning, resources and costs, and partners), activities and outputs of the “10,000 Lives” initiative of Central Queensland, Australia.

## Method

An evaluation plan was formulated to investigate the inputs, activites, outputs, impact and outcome of the “10,000 Lives” initiative. A program logic model, adapted from a standard health promotion evaluation framework,^19^ was developed for the evaluation plan. The evaluation framework was discussed among stakeholders who attended the “10,000 Lives” summit in Rockhampton, Australia, in November 2018. The model has guided understanding the program inputs and outputs for the evaluation. The model, shown in **Figure 1**, demonstrates the process evaluation framework by illustrating the interplay of the different factors that may influence the impacts and outcomes of the program activities. However, this paper focuses on describing the inputs (planning, resources and cost, and partnership), activities and outputs of the initiative.

**Figure 1.**
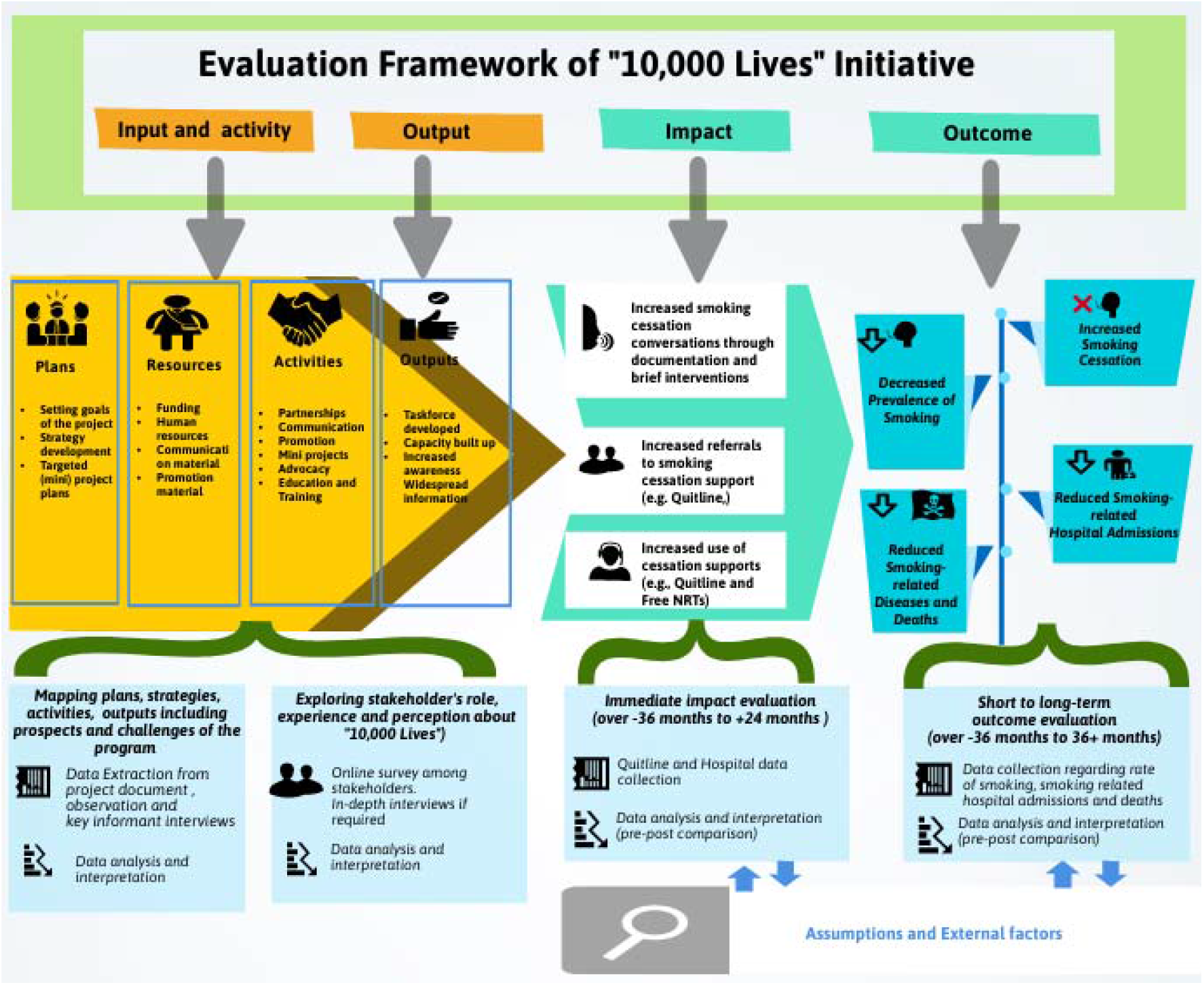
Logic model for evaluation of “10,000 Lives” initiative

### Target population of the “10,000 Lives” initiative

The target population for the “10,000 Lives” initiative is all smokers living in the service catchment area of CQHHS (**Figure 2)**, which includes 12 public hospitals in CQ.^5^ In 2017, the population of the CQ region was ∼220,000 people (4.5% of the Queensland population and 0.9% of the Australia population).^20^ There were 54,722 families (74,201 households) in 2017; the median age was 34.9 years; sixty-five per cent of the population were aged between 15–64 years.^21^ Approximately six per cent of the population are Aboriginal and/or Torres Strait Islanders.^7^ The rate of homelessness was 41.0 per 10,000 persons. The median total personal income per year was $35,017 Australian dollars (AUD), with 50.2% having the highest level of schooling of Year 11 or 12 (or equivalent). In CQ, 25.7% of the population were in the most disadvantaged quintile and 10.1% of the population were in the least disadvantaged quintile, whereas in Queensland, 20% of the population were in most disadvantaged quintile and 20.0% in the least disadvantaged quintile in 2017.^7^ According to a state-wide survey in the year preceding the launching of “10,000 Lives” an estimated ∼28,000 adult daily smokers resided in CQ.^22^ The daily smoking prevalence was highest (17.4%) in the 30–44 years age group. Also, the prevalence was high (18.5%) among the most disadvantaged quintile. The proportion of the determinants for poor health, i.e., ‘low-income households’, ‘early exit from school’, ‘unemployment’ and ‘mental health issue’ is higher in CQ than the whole of Queensland and Australia.^23^

**Figure 2.**
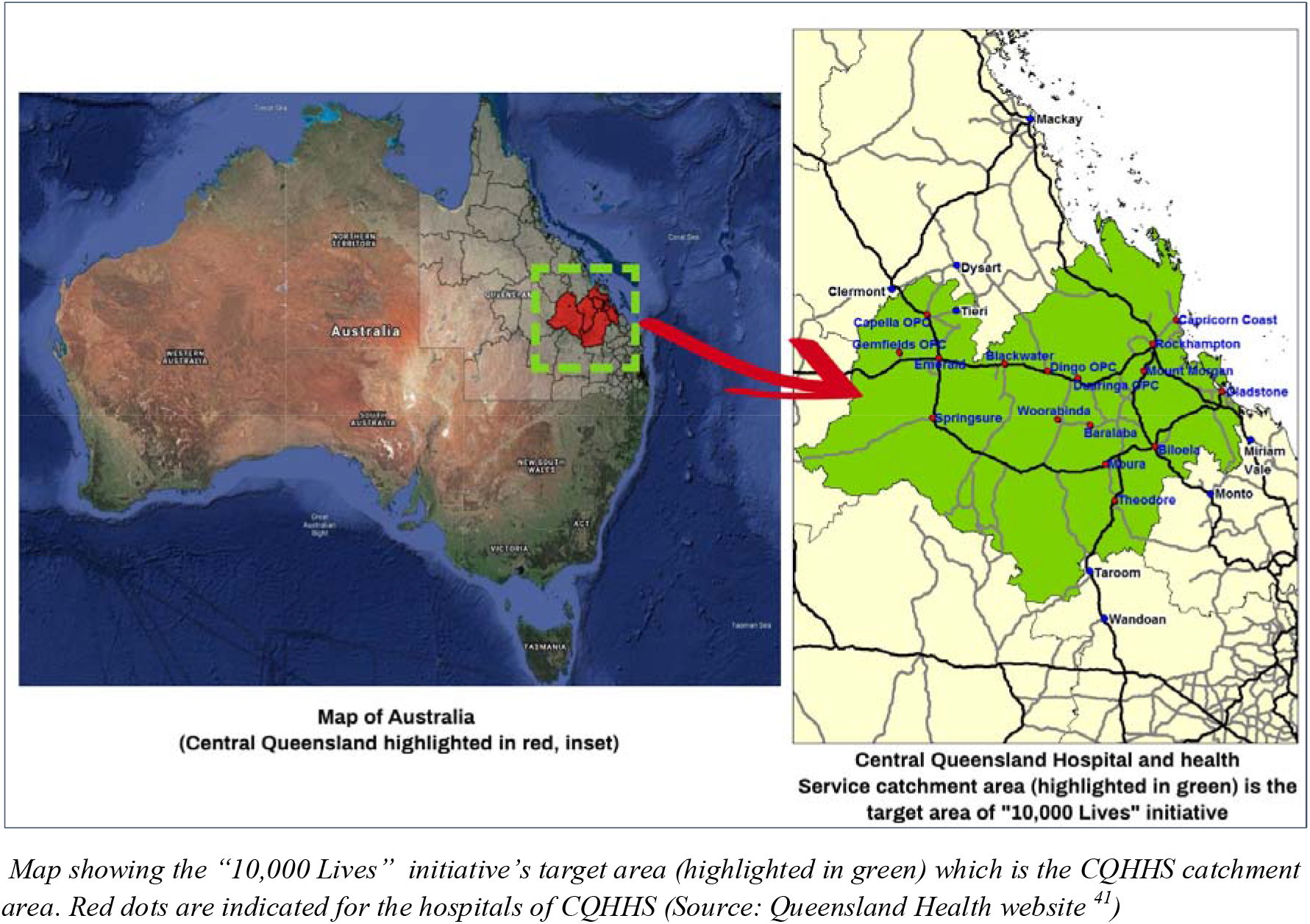
Study Area Map; “10,000 Lives” initiative’s catchment area

### Study design, data collection and analysis

We conducted an exploratory investigation by critically appraising the project plan, partnership development, communication strategies, targeted project activities and overall health promotion activities for smoking cessation covered by the “10,000 Lives” initiative. Data were collected retrospectively for the period between July 2017 (initiation of planning) up to December 2019 (26 months after the official launch of “10,000 Lives”) from field notes, project documentation notes, relevant policy documents, and key informant interviews with project personnel.

A generic search was performed of relevant websites (i.e., Queensland Health: http://www.health.qld.gov.au, CQ Health: http://www.health.qld.gov.au/cq, Department of Health of Australian Government: http://www.health.gov.au, and Australian Institute of Health and Welfare: http://www.aihw.gov.au) for relevant policy documents, and social media pages (e.g., Facebook) for information about smoking cessation campaigns active during the study timeframe.

Data were extracted from these sources and imported into NVIVO,^24^ and then cleaned, coded, and classified into five themes: plans, resources and cost, partnerships, activities and outputs. A narrative synthesis and summary interpretation was completed and these are presented in the results section. The data sources and collection methods are shown in **Table 1**.

**Table 1.** Summary of data sources and collection method for each evaluation topic

| Themes | Data sources | Collection method |
| --- | --- | --- |
| *Inputs* |  |  |
| Planning | Project planning documents and policy document ‘Destination 2030’<sup>13</sup> | Document review and internet search |
| Resource | Project management documents | Document review |
| Partnership | Project planning and management documents, Key informant interviews | Document review, Key informant interviews |
| *Activities* |  |  |
|  | Project management documents, Master file for project management, Key informant interviews, Working group documents, Websites and Social media | Document and content review, Websites, Social media, Observation, Key informant interviews |
| *Outputs* |  |  |
|  | Project management documents, Master spread sheet, Stakeholders meeting documents, Attendance sheet, and Key informant interviews | Document review, Key informant interviews |
| *Anticipated impact and outcome* |  |  |
|  | Project management documents, Policy documents, Key informant interviews | Project document review, Policy documents review, Key informant interviews |

### Ethical approval

The study was approved by CQHHS Human Research Ethics Committee (HREC) (HREC/2019/QCQ/50602).

### Patient and Public involvement

We used routine data source for process evaluation of the program. Individual participants were not involved in this study.

## Result

**Table 2** lists the key findings of the important areas for process evaluation (i.e., planning, resources and cost, partnerships, activities and project outputs) covered in this study in the first 26 months since the “10,000 Lives” initiative launched. Major strategies employed by the “10,000 Lives” initiative are shown in **Figure 3**.

**Table 2.**
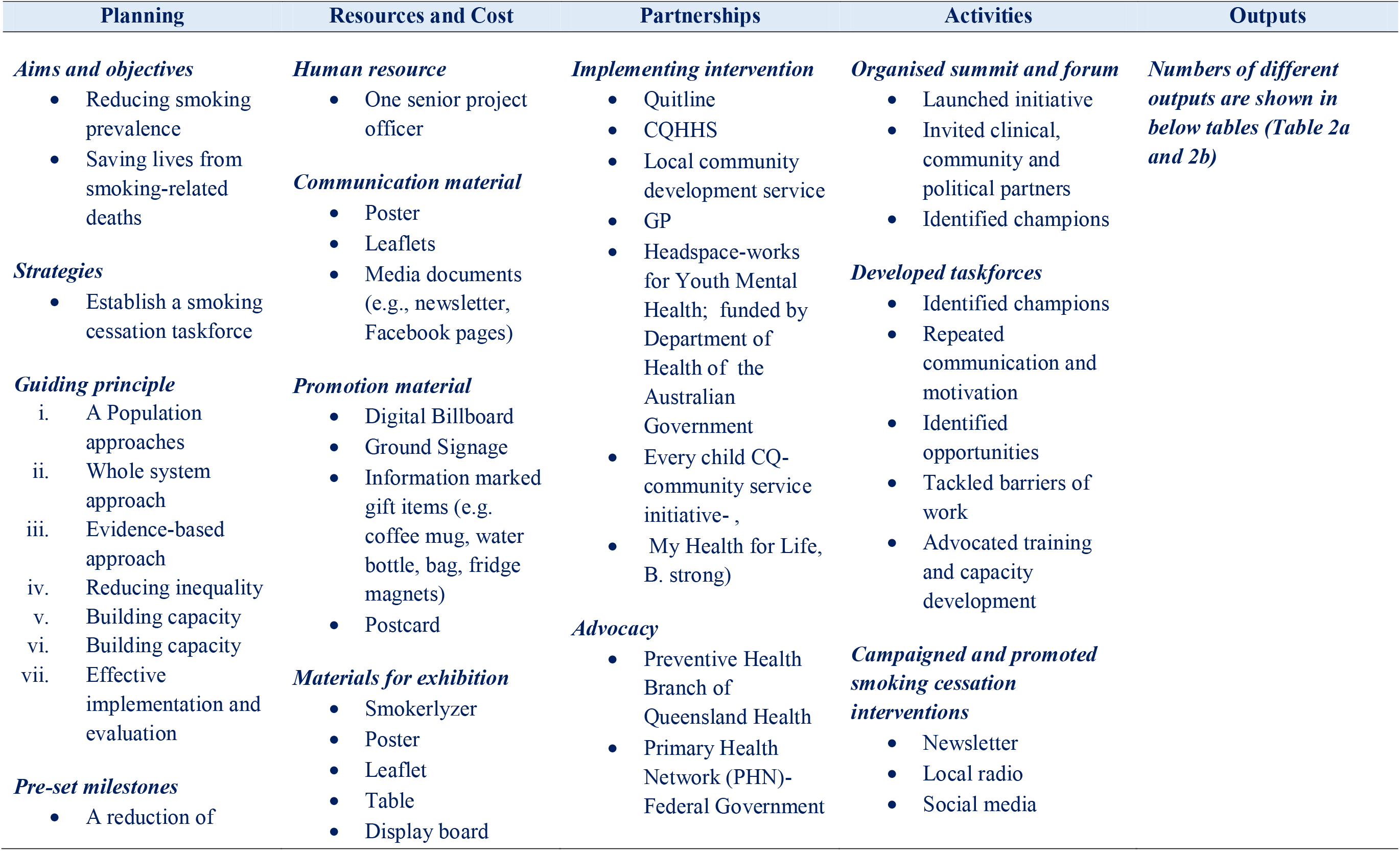
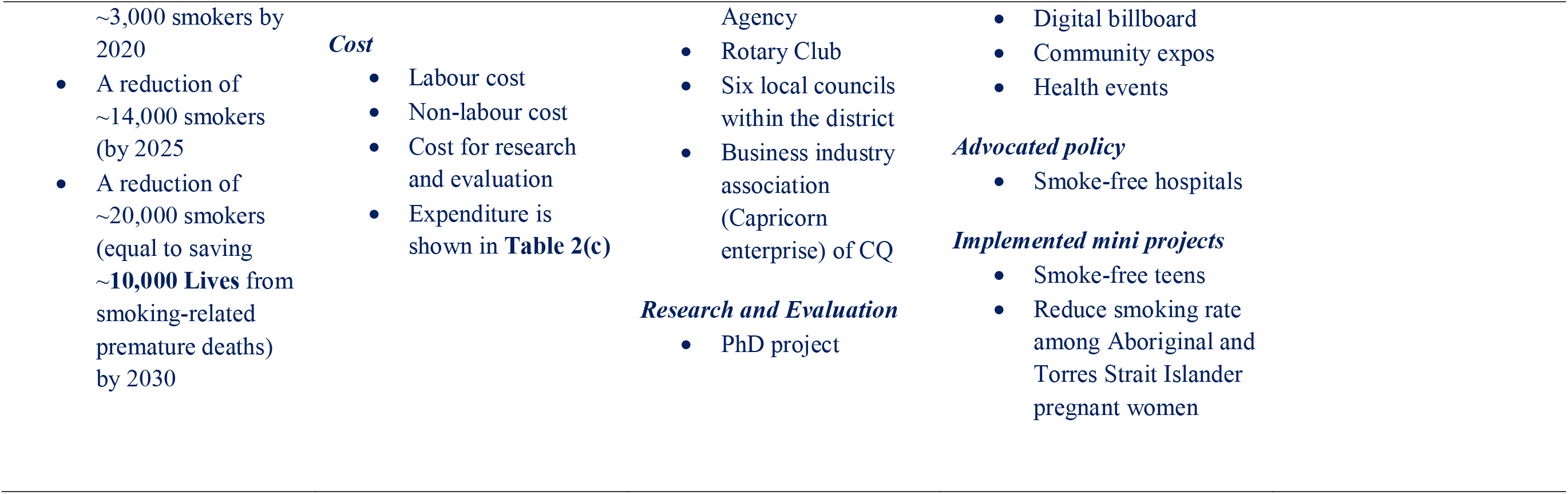
Description of project planning, resources, partnerships, activities and outputs in first 26 months of project launched

| Planning | Resources and Cost | Partnerships | Activities | Outputs |
| --- | --- | --- | --- | --- |
| *Aims and objectives*<br>Reducing smoking prevalence<br>Saving lives from smoking-related deaths<br>*Strategies*<br>Establish a smoking cessation taskforce<br>*Guiding principle*<br>A Population approaches<br>Whole system approach<br>Evidence-based approach<br>Reducing inequality<br>Building capacity<br>Building capacity<br>Effective implementation and evaluation<br>*Pre-set milestones*<br>A reduction of | *Human resource*<br>One senior project officer<br>*Communication material*<br>Poster<br>Leaflets<br>Media documents (e.g., newsletter, Facebook pages)<br>*Promotion material*<br>Digital Billboard<br>Ground Signage<br>Information marked gift items (e.g. coffee mug, water bottle, bag, fridge magnets)<br>Postcard<br>*Materials for exhibition*<br>Smokerlyzer<br>Poster<br>Leaflet<br>Table<br>Display board | *Implementing intervention*<br>Quitline<br>CQHHS<br>Local community development service<br>GP<br>Headspace-works for Youth Mental Health; funded by Department of Health of the Australian Government<br>Every child CQ-community service initiative- ,<br>My Health for Life, B. strong)<br>*Advocacy*<br>Preventive Health Branch of Queensland Health<br>Primary Health Network (PHN)- Federal Government | *Organised summit and forum*<br>Launched initiative<br>Invited clinical, community and political partners<br>Identified champions<br>*Developed taskforces*<br>Identified champions<br>Repeated communication and motivation<br>Identified opportunities<br>Tackled barriers of work<br>Advocated training and capacity development<br>*Campaigned and promoted smoking cessation interventions*<br>Newsletter<br>Local radio<br>Social media | *Numbers of different outputs are shown in below tables (Table 2a and 2b)* |

|  |  |  |  |
| --- | --- | --- | --- |
| ~3,000 smokers by 2020<br>• A reduction of ~14,000 smokers (by 2025)<br>• A reduction of ~20,000 smokers (equal to saving ~10,000 Lives from smoking-related premature deaths) by 2030 | *Cost*<br>• Labour cost<br>• Non-labour cost<br>• Cost for research and evaluation<br>• Expenditure is shown in **Table 2(c)** | Agency<br>• Rotary Club<br>• Six local councils within the district<br>• Business industry association (Capricorn enterprise) of CQ | • Digital billboard<br>• Community expos<br>• Health events |
|  |  | *Research and Evaluation*<br>• PhD project | *Advocated policy*<br>• Smoke-free hospitals |
|  |  |  | *Implemented mini projects*<br>• Smoke-free teens<br>• Reduce smoking rate among Aboriginal and Torres Strait Islander pregnant women |

**Table 2(a).** Occurrence of different activities and events implemented by “10,000 Lives” initiative in first 26 months

| Event name | Frequency (n) |
| --- | --- |
| Smoking cessation summits organised | 7 |
| ‘World No Tobacco Day’ campaign (including a tobacco forum) | 2 |
| Number of mini-projects implemented for priority populations | 2 |
| Film competition with the theme of smoking cessation for young people, organised | 1 |
| Attended health event with “10,000 Lives” stall | 9 |
| Attended community expo and event with "10,000 Lives stall" | 12 |
| Brief education sessions delivered to newly recruited CQHHS staff | 20 |
| In-person meeting conducted | 97 |
| Facebook pages (10000 Lives CQ, CQ Health and Triple M central Queensland) discussed posts | 206 |
| Occasional share of the updated information and resources to CQHHS staff through daily news and the weekly bulletin (Drift) | 4,800 |
| Occasional emails with updated information and resources to the personnel from partner organisations other than CQHHS | 3,409 |

**Table 2(b).**
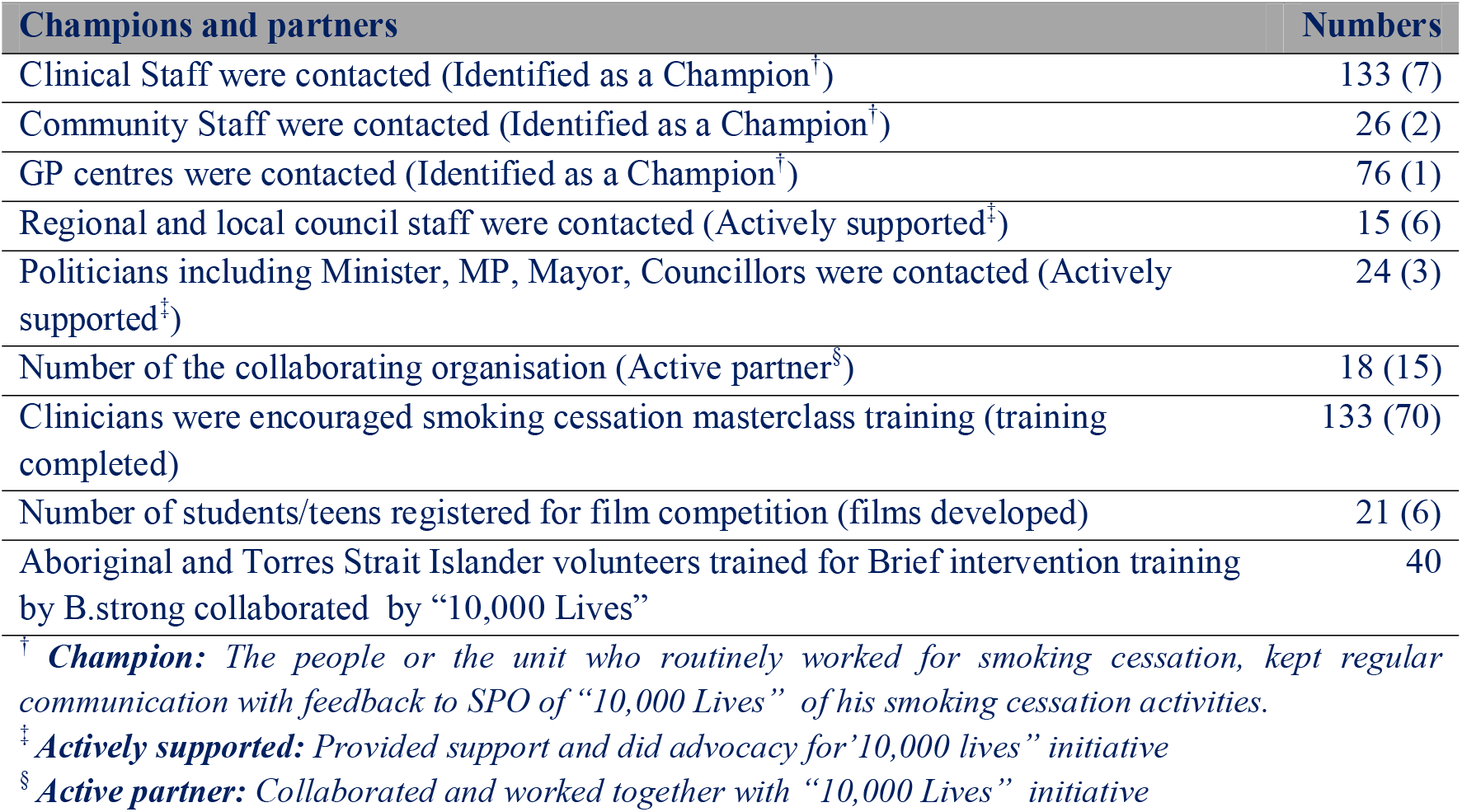
Numbers of partners and champions contacted, and numbers who supported the “10,000 Lives” initiative in first 26 months

**Table 2(c).** Monetary cost spent for “10,000 Lives” initiative in 24 months (January 2018-December 2019) after launch

| Item of cost | Amount in Australian Dollar |
| --- | --- |
| Labour cost (human resource) | \$199,600.0 |
| Non-labour cost (Materials, supplies, travel etc.) | \$16,984.0 |
| Evaluation and research cost (PhD project) | \$64,164.0 |
| *Total* | *\$ 280,748.0* |

**Figure 3.**
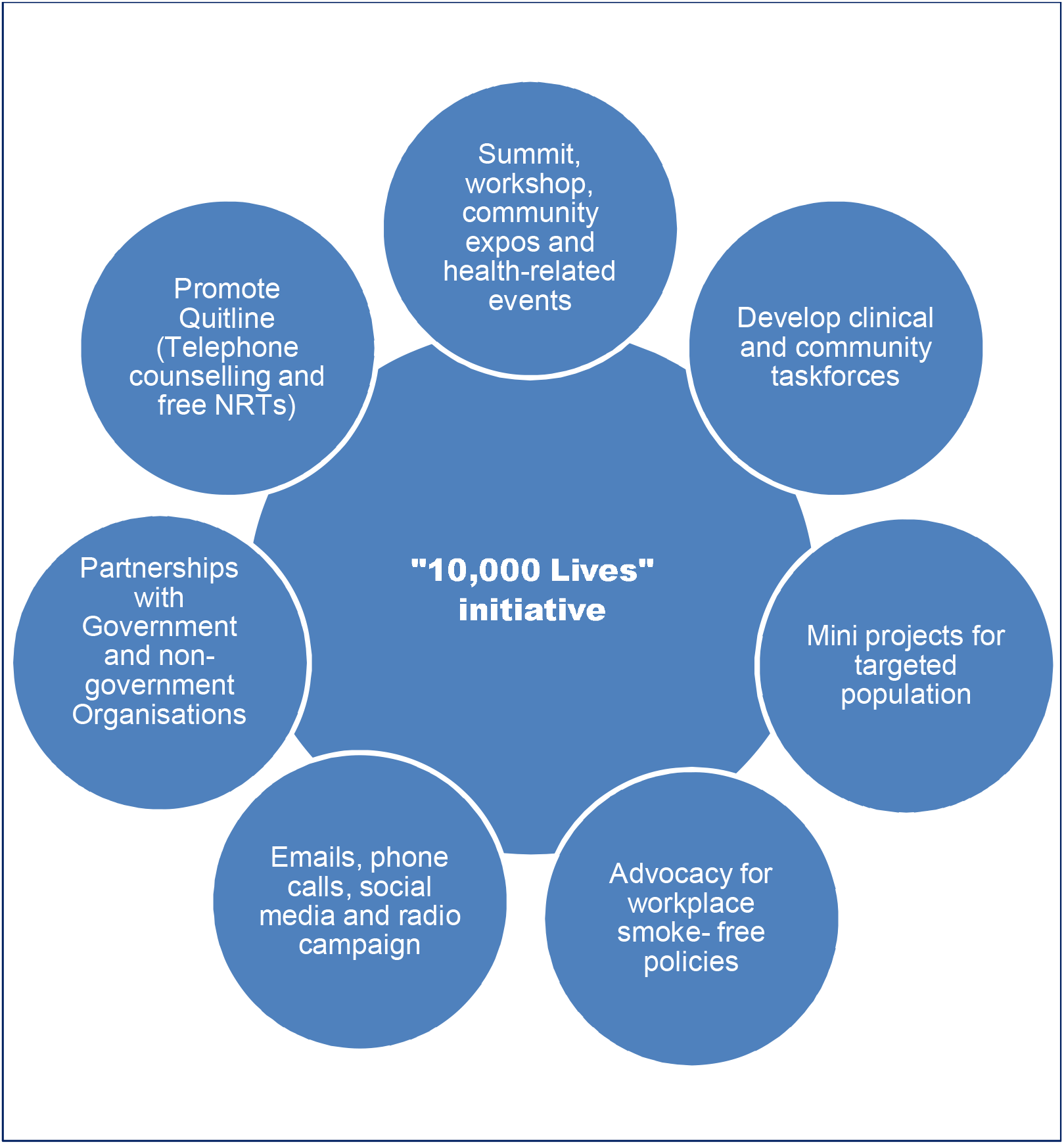
Major strategies employed by “10,000 Lives” initiative

### Planning

At the program planning stage (July-August 2017), CQPHU, with the help of the Service Integration Coordinator of the Department of CQ Mental Health Alcohol and Other Drugs, developed a project proposal to establish a smoking cessation taskforce in CQ. The project proposal^25^ stated the objectives of the initiative as:

1. *Establish a 10,000 Lives Taskforce: The taskforce will form the backbone of the project and through collective impact with the support of a wide range of community stakeholders large scale social change will be achieved. (“Collective impact” is a structured and disciplined approach to bringing cross-sector organisations together to focus on a common agenda that result in long-lasting improvement.)*.
2. *Establish a team of clinical champions to engage key stakeholders e.g. G.P.’s and provide health promotion activities, intervention and education to the broader community”*.

The aim was subsequently reflected in ‘Destination 2030’.^13^ The initial plan considered strategies that adhered to the following guiding principles: i) *Population approach* of delivering a sustained, effective and comprehensive initiative for all, ii) *Whole system approach* of harnessing the many inter-related factors that can contribute to improving health and wellbeing, iii) *Evidence-based approach* of integrating knowledge from research evidence into implementation, iv) *Reducing inequality* by addressing the differences in health status in the community through recognising and responding to the vulnerable groups (e.g. the groups who have higher smoking prevalence), v) *Working in partnership* with government departments, community members, NGOs, and academic stakeholders, vi) *Building capacity* by developing an adequate number of skilled and empowered people, and vii) *Effective implementation and evaluation* for ensuring the platform to track the collective impact.^25^ For implementing the approaches, multiple and specific mini-projects were planned to target priority groups. For example, plans were formulated to give more attention to specific geographical areas (e.g., Gladstone and Woorabinda) and populations (e.g., mine workers and Aboriginal and Torres Strait Islander people). Ambitious milestones were set during program planning including a reduction of ∼3,000 smokers by 2020’, ‘a reduction of ∼14,000 smokers by 2025’ and ‘a reduction of ∼20,000 smokers by 2030’.^25^

### Resources and costs

A senior project officer (SPO, Administrative Officer Grade 5) was recruited in December 2017 to coordinate the planned activities and manage the implementation of the program strategies. Other resources utilised in the project included communication materials (e.g., Posters and leaflets, emails, news, website and social media content), promotion materials (e.g., information containing postcards, coffee cups, fridge magnets, water bottles and bags), materials required for mobile stalls to display the project activities in community or health events (e.g., display table, carbon monoxide breath testing for smokers), organising summits and workshops, and ground signage. The approximate cost for running the program for 24 months (January 2018-December 2019) was $280,748 (AUD) including the amount $64,164 (AUD) for the research and evaluation component **(Table 2(c))**. The initiative was approved by the CQHHS board and solely funded by CQHHS.

In addition to the direct resource and costs, the initiative utilised in- kind support from the CQPHU for administrative activities including administration staff support and operational support during the period between starting the program planning in July 2017 and the official launch of the initiative in November 2017. Also, the initiative utilised the existing resources available for smoking cessation in CQ which included combination of 12-weeks-free NRTs and telephone counselling via the Queensland Quitline’s intensive Quit support program,^12^ subsidised smoking cessation pharmacotherapies through the Pharmaceutical Benefits Scheme, Queensland Health’s Quality Improvement Payment (an incentive program for clinicians), and the collaborative support from existing smoking cessation programs (i.e., “Quit for You…Quit for Baby”, “Quit for You”, “Yarn to Quit”, B.strong).

### Partnerships

Developing partnerships and involving stakeholders in the implementation of “10,000 Lives” was a key strategy of the initiative. A strategic partnership was made with the Queensland Quitline^11^ for enhancing the promotion of their existing intensive Quit support program which was available to rural, regional and remote communities with a higher than average smoking prevalence and accessing a monthly report to track Quitline registrations and participation status for smokers in CQ. Extensive in-kind support was provided by the Board and Chief Executive of CQHHS by arranging the project fund, and the Preventive Health Branch of Queensland Health by giving strategic advice and advocacy for implementing the smoke-free policies. Partnerships were built with different units and programs within CQHHS (e.g., Oral health, Mental health, ‘CQ Youth Connect’), community organisations (e.g., Rotary^26^), a foundation for youth mental health called ‘Headspace’,^27^ a targeted brief intervention training program for Aboriginal and Torres Strait Islander people named ‘B.strong’,^28^ a health promotion initiative for Aboriginal and Torres Strait Islander people called ‘Deadly Choices’,^29^ local councils (city council and local government staff) and a non-government organisation (NGO) supporting and developing buisnesses and projects in CQ called “Capricorn Enterprise”^30^ to promote and support smoking cessation activities for their own staff and client population (patient, youth, community and Aboriginal and Torres Strait Islander people who smoke). The project collaborated with a University in Australia for academic support for the program evaluation. Partnerships were developed with “Cancer Council Queensland”^31^ for conducting training and workshops for the local clinicians, social workers and volunteers who were interested in supporting the initiative. The local Primary Health Network (PHN) actively collaboratored with “10,000 Lives” initiative by distributing information to General Practitioners (GPs). Local sports clubs and radio staions also partnered with the initiative on health promotion activities.

### Activities

The SPO coordinated the activities of “10,000 Lives” under the guidance of the director of CQPHU. The SPO took a pre-set plan and continuously adapted strategies (described in planning section) for implementing the program. The following range of activities were delivered to increase smoking cessation in CQ:

1. **Organising tobacco summits** to develop partnerships with clinicians, GPs, social workers, local council and industry staff, and local politicians.
2. **Establishing a clinical and community organisation taskforce** for smoking cessation to identify clinical and community organisation personnel to become a champion for smoking cessation. CQHHS clinicians were encouraged to conduct inpatient hospital and health care facility-based documentation and brief intervention via a standardised ‘Smoking Cessation Clinical Pathway (SCCP)’ form among patients who smoke, and to refer them to Quitline for accessing the intensive Quit support program. Community champions were encouraged to promote the Quitline program and other smoking cessation support (e.g., My QuitBuddy app) among people who smoke.
3. **Promoting smoking cessation** through emails, newsletters, local radio, social media pages (i.e., Facebook), digital billboard and ground signage, and exhibiting in various community expos and health-related events. The SPO explored various communication pathways to promote the available smoking cessation support, particularly the Quitline program. These included; conducting events on the local radio station (‘Triple M’), posting messages on Facebook pages (“10,000 Lives”, CQHHS and ‘Triple M’ Facebook pages), local newspapers (The Morning Bulletin and Gladstone Observer) and in the daily news and weekly bulletin of CQHHS and e-newsletters for GPs, and electronic billboard display in the center of the main city of the CQ region (i.e. Rockhampton CBD).
4. **Advocating for smoke-free policies and programs** that could support smokers to quit. For example, the initiative established the ground signage and delivered tear off flyers promoting Smoke-free Healthcare in each of the hospital and community health campuses of CQHHS.
5. **Implementing mini-projects** to give extra attention to priority populations. For example, a film competition on ‘smoke-free teens’ was organised to deliver a youth-centric smoking cessation message designed by youth for youth, and a workshop was conducted by the SPO to introduce carbon monoxide breath testers (**Smokerlyzer)** with *Gumma Gundoo Indigenous Maternal & Infant Care Outreach team*^32^ to increase awareness amongst Aboriginal and Torres Strait Islander pregnant women of the adverse effects of antenatal smoking on mother and baby.

### Outputs

The quantitative output measures from the “10,000 Lives” activities are shown in Table 2(a) and 2(b). Overall, the “10,000 Lives” initiative conducted seven smoking cessation summits and one Tackling Tobacco Forum, promoted and celebrated World No Tobacco Day regionally, completed at least twenty education sessions for newly recruited CQHHS staff, and conducted a combined smoking cessation workshop for the clinical and community champions. The SPO encouraged all the clinicians of CQHHS to attend the Smoking Cessation Masterclasses conducted by Queensland Health (Metro South HHS and Metro North HHS), with 70 clinicians completing, and a three-day training course on nicotine addiction and smoking cessation,^33^ which was completed by six clinicians. Forty Aboriginal and Torres Strait Islander volunteers were trained in Brief Intervention training conducted by the Menzies School of Health Research (B.strong).^28^ The “10,000 Lives” initiative was exhibited in twelve community expos and nine health-related events. The initiative implemented two mini-projects for priority population (Aboriginal and Torres Strait Islander pregnant women, younger people). “10,000 Lives” collaborated with 15 different organisations including Hospital and Health Service, regional councils, University, Community Organisations and other initiatives to promote smoking cessation in CQ. The SPO shared updated resources and information about smoking cessation to ∼3,400 staff of different partner organisations through emails and 4,800 staff of CQHHS through posting in the daily news and a weekly bulletin called ‘The Drift’. As a result of communication through email, phone call, posting messages and in-person meetings by the SPO, at least seven clinical champions, two community champions, two political champions and a champion GP centre became actively involved and worked on the ground as the smoking cessation taskforce in CQ.

## Discussion

This study describes the inputs, activities and outputs of the program logic model, documenting the process evaluation of the “10,000 Lives” initiative. This article explains why and how the initiative was implemented, and describes the way it operated over the 26 month period following its official launch in November 2017. This study also outlines how success of the program will be measured.

The “10,000 Lives” initiative was launched to reduce the daily smoking rate in CQ, which is higher than the state average. Policymakers realised the high disease burden that is due to smoking and included the ambitious aim to reduce the smoking rate to 9.5% by 2030 in the Destination 2030 plan. The implementing organisation of the initiative is the local Public Health Unit which explored the existing and available smoking cessation support available in its region. A number of effective tobacco control and smoking cessation interventions were already available in the region, and the “10,000 Lives” initiative aimed to increase awareness and uptake of these interventions. In this way, the initiative focused on maximising the use of existing services available in the region.

The initial plan was guided by the standard principles (described in the *Planning* part of the results section) of program implementation. The initiative was launched in each of the local government areas of CQ region at a Smoking Cessation Summit. People from multiple sectors including Health and Community Services and state and local government were invited to attend the inaugural summit which ultimately facilitated the initiative to build the partnerships and identify champions. Partnerships were built with various government and non-government organisations so that the coverage of workplace based smoking cessation programs were increased and the smoke-free workplace policies implemented. Active partnership with Quitline Queensland assisted the initiative to promote their intensive Quit support program. The SPO was integral to building communication pathways to promote the smoking cessation support available. The regular communication and motivation to the stakeholders (clinical and community champions) helped the SPO to identify the opportunities (e.g., arrange training and workshop on smoking cessation) and tackle the barriers of smoking cessation work for them. This integral strategy of building partnerships and communication pathways became useful to build a clinical and a community taskforce of smoking cessation in CQ which leveraged the existing smoking cessation program and policies available in the region. This approach is an exemplar of running a health promotion campaign in a resource constrained environment.

The 10,000 “Lives” initiative was built on the success of a previous health promotion campaign “10,000 Steps Rockhampton” in this region.^16,17^ The Rockhampton area was choosen for “10,000 Steps Rockhampton” program because of the high prevalence of obesity.^16^ Again, “10,000 Lives” initiative was launched in CQ to address the higher prevalence of smoking in this region. The “10,000 Lives” utilised the program strategies (e.g., media campaign, partnerships with clinicians, focusing on priority populations) that were also used in the “10,000 Steps Rockhampton” program.^16^ Other similarities include the use of of technology to measure exhaled carbon monoxide in “10,000 Lives” and pedometers in “10,000 Steps Rockhampton” to measure activity levels. The use of the Smokelyzer provided a teaching moment to discuss the health impacts of smoking by demonstrating the person’s exposure to one of the toxins in cigarette smoke, leading to increased autonomous-motivation to quit smoking. Creating autonomus motivation in people who smoke, often explained by the ‘Self-Determination Theory’,^34^ is effective for promoting smoking cessation.^35^

However, the implementation of the program was sometimes challenging, such as increasing clinician participation in deliverying brief advice and quitline referrals. Some stakeholders expected “10,000 Lives” to directly deliver smoking cessation services. However, this was beyond the scope and resources of the program.

The “10,000 Lives” initiative is quite different from other smoking cessation programs in Australia (e.g., B.strong, Quitline) which deliver smoking cessation assistance directly to smokers. Rather, “10,000 Lives” intended to increase motivation to quit, and raise awareness of existing smoking cessation assistance that is available via these other programs. While the national tobacco campaign^36^ and statewide anti-smoking campaigns primarily use paid advertising to disseminate the quit smoking message, the “10,000 Lives” program focused on low cost approaches to disseminating the quit smoking message via partnerships with local media and local clinical and community champions for promoting the smoking cessation interventions. This model has also been used in other health promotion programs implemented in New South Wales, Australia and in a community of North East England.^37,38^ However, the findings of process evaluation using a logic model of those program were not found after serachin in relevant websites.

The strategies for achieving the goal of the “10,000 Lives” initiative reflect ecological models of health promotion’ which explain the multiple levels of influence on health behaviour.^39^ The initiative put substantial efforts to increase the use of interventions of smoking cessation programs by involving the service providers in the community (e.g., clinicians, NGO personnel) such this is a ‘downstream’ approach. For example, the “10,000 Lives” initiative encouraged clinicians to deliver brief interventions with their patients and refer them to Quitline, and other relevant smoking cessation programs. The use of local radio, which involved sports stars discussing smoking cessation and posting messages on Facebook pages are examples of ‘midstream’ strategies. While the advocacy of state level policies and programs (e.g., smoke-free hospitals) are ‘upstream’ strategies. Thus the “10,000 Lives” program fits the multi-level population based health promotion model of McKinlay.^40^

Overall, the initiative brought all the available smoking cessation support together and promoted the adoption of smoke free policies and programs. The stakeholders’s perspective and the impact of “10,000 Lives” are currently evaluated through analysis of the stakeholder-survey data and changes in the numbers and rate of referrals, program participation and interactions to Quitline, and the result will be reported through separate peer-reviewed publications.

## Conclusion

The “10,000 Lives” is an example of a health promotion program which coordinates smoking cessation activities in a regional area by harnessing and improving awareness of existing resources (e.g. employing only one project officer). Utilising existing resources and programs can be a cost-effective approach in countries like Australia where effective smoking cessation interventions are already widely available, but uptake is suboptimal. Evaluation of impact and outcome of this initiative could inform the development of future regional smoking cessation programs.

## Data Availability

Not applicable

## Contributors

AK, GK, SL and CG conceived and designed the study. AK conducted the key informant interviews. AK and KG extracted the data from different sources. AK performed the analysis of the data. AK, KG, GK, SL and CG interpreted the results. AK drafted the manuscript and all authors contributed with critical revisions to the contents of the manuscript. The final version of the manuscript was approved by all authors.

## Acknowledgement

We acknowledge the unrestricted support from Central Queensland Public Health Unit, Central Queensland Hospital and Health Service, Preventive Health Branch and Quitline Services from Queensland Health (Queensland Government). We thank Caron Williams, the first senior project officer of the “10,000 Lives” program, and Susie Cameron, the service integration officer of CQ Mental Health Alcohol and Other Drugs, for sharing the project information through informal discussions and emails. Also, we thank Linda Medlin, Acting Director of Aboriginal and Torres Strait Islander Health and Wellbeing in Central Queensland Hospital and Health Service for her kind review of the manuscript in the perspective of cultural respectability.

